# Test Sensitivity for Infection versus Infectiousness of SARS-CoV-2

**DOI:** 10.1101/2020.08.30.20184739

**Authors:** Joshua S. Gans

**Affiliations:** Rotman School of Management, University of Toronto and NBER

**Author notes:** All correspondence to. Disclaimer: I am an economist and not an epidemiologist. I have received no funding for this research and have no conflicts of interest. Thanks to Laura Rosella, Jakub Steiner and Alex Tabarrok for useful comments. Responsibility for all views expressed and errors made lies with the author.

## Abstract

The most commonly used test for the presence of SARS-CoV-2 is a PCR test that is able to detect very low viral loads and inform on treatment decisions. Medical research has confirmed that many individuals might be infected with SARS-CoV-2 but not infectious. Knowing whether an individual is infectious is the critical piece of information for a decision to isolate an individual or not. This paper examines the value of different tests from an information-theoretic approach and shows that applying treatment-based approval standards for tests for infection will lower the value of those tests and likely causes decisions based on them to have too many false positives (i.e., individuals isolated who are not infectious). The conclusion is that test scoring be tailored to the decision being made.

## 1 Introduction

An intuitive notion that guides tests for the presence of a virus in an individual is that it is preferable to have tests that have the capability to detect smaller loads of the virus in any given sample (e.g., blood, saliva or nasal mucus). As the presence of the virus is a necessary condition for someone to be infectious – that is, to have a positive probability of transmitting the virus to susceptible person – medical practitioners and government regulators often set standards for a minimum amount of a virus that a test needs to be capable of identifying before or using that test for clinical purposes. However, while being infected is a necessary condition for infectiousness, it is not sufficient. With the Covid-19 pandemic of 2020, it has been discovered that individuals who are infected – in terms of having severe acute respiratory syndrome coronavirus 2 (SARS-CoV-2) present – may not be infectious. This is because infectiousness both requires an individual to have a sufficient viral load and the virus present has to be active. This implies that, if your relevant clinical decision is to isolate an individual to prevent infections in others, as this paper will show, the intuition that you prefer a more precise test falters and less precise tests can be more valuable.^1^

The primary means of testing for SARS-CoV-2 is a reverse transcriptase-quantitative polymerase chain reaction (RT-qPCR) test. Such PCR tests use a technique (PCR) to test for viral RNA remnants in cycles where RNA segments are exponentially replicated in order to increase the likelihood of even small numbers of them being identified in a sample. The test stops once the targeted RNA is identified or, typically, after 40 cycles. If the test run completes without the RNA being found, the test result is returned as ‘negative.’ Otherwise, it is ‘positive’ and the individual is held to be infected. This process requires a laboratory, reagents and specialised machines and can cost between $50-150 per test and take between 24 and 48 hours for results to be returned.

The cost of PCR tests, along with the length of time taken for results to be communicated to medical practitioners, has led to calls for cheaper, rapid tests to be used in order to mitigate the spread of Covid-19 (the disease caused by SARS-CoV-2).^2^ Larremore et al. (2020) note that a typical PCR test can detect the virus up to 10 copies per million (cp/ml) while point-of-care nucleic acid LAMP or rapid antigen tests can only detect up to 10^5^ cp/ml. These tests are not to be as accurate as PCR tests for small viral loads but it is also noted that the threshold for infectiousness is more likely 10^6^ cp/ml. Importantly, Larremore et al. (2020) note that even if an infected patient is caught at 10 cp/ml, the time it takes for their load to increase above 10 cp/ml is short and may be negative once the time taken to process a PCR test is taken into account. Moreover, after the most infectious period in an individual, the PCR tests can still detect infections and, indeed, can detect viral remnants that may not be alive.

The typical path of the viral load for SARS-CoV-2 is shown in Figure 1.^3^ Suppose that a PCR test takes 48 hours to return a result. Then if that test is taken at Day 3 (point A) then the result will be returned on Day 5 (post C) when the individual has potentially been infectious for a day. By contrast, an antigen test taken on Day 3 would return a negative result but if it were used daily and taken also on Day 4 (point B), that individual would be positive and could be isolated immediately. Thus, even though the antigen test is less accurate for identifying an infection than PCR, its cost and consequently frequency of application that allows may make it a more effective tool for mitigating the spread of Covid-19.^4^ Larremore et al. (2020) conclude that “the FDA, other agencies, or state governments, encourage the development and use of alternative faster and lower cost tests for surveillance purposes, *even if they have poorer limits of detection.”* (p.7, emphasis added)

**Figure 1:**
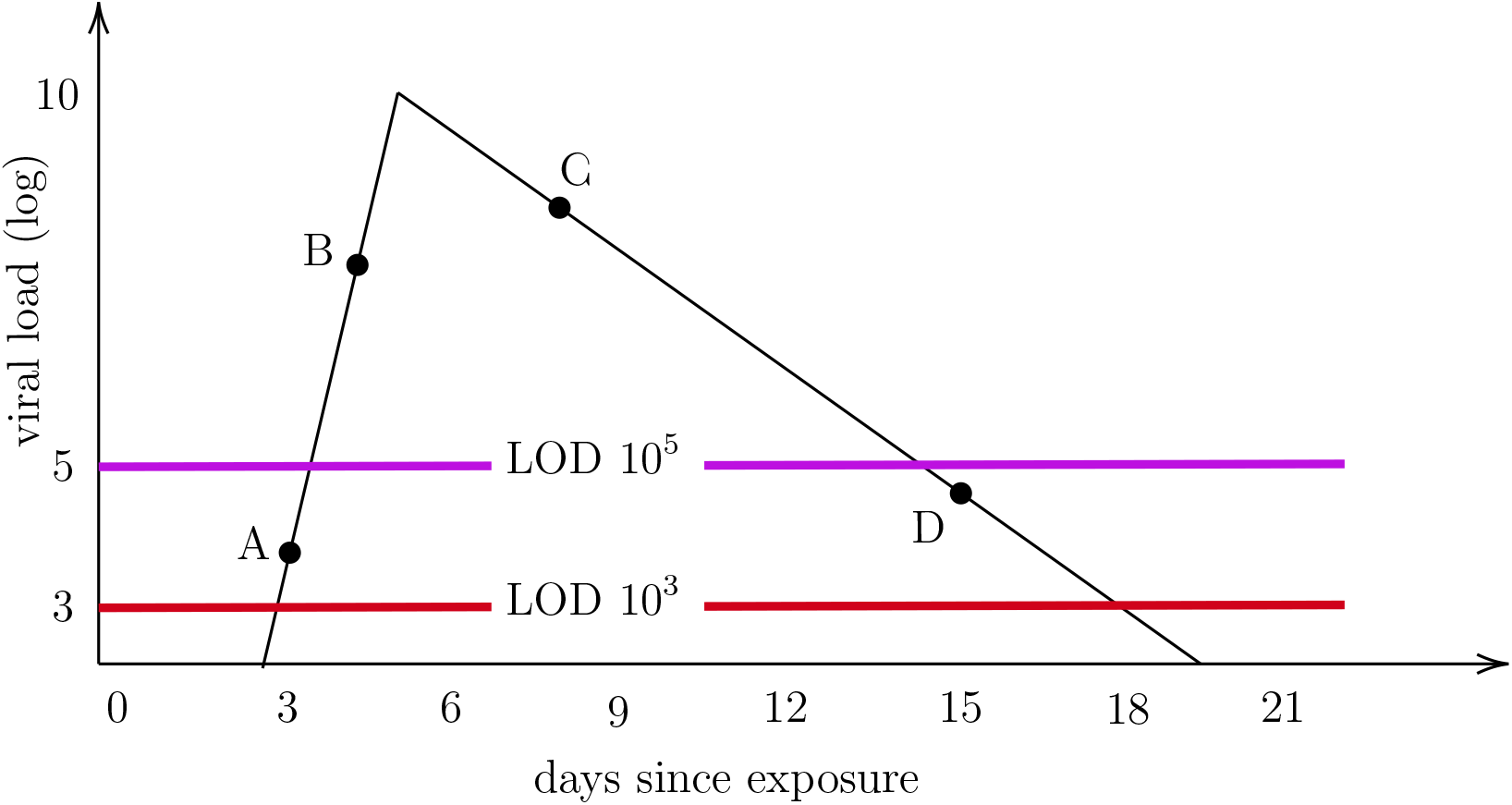
Viral Load of Infected Individual Over Time

**Figure 2:**
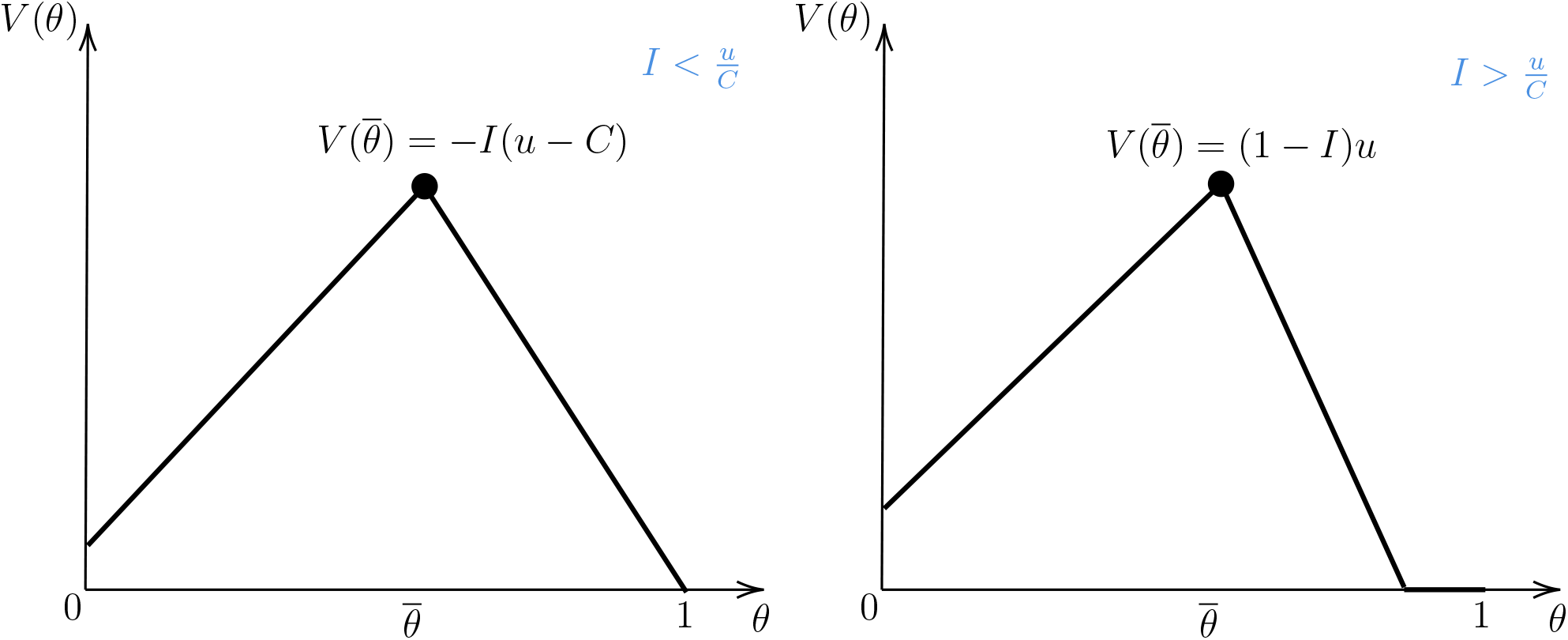
Value of Tests for Infectiousness

**Figure 3:**
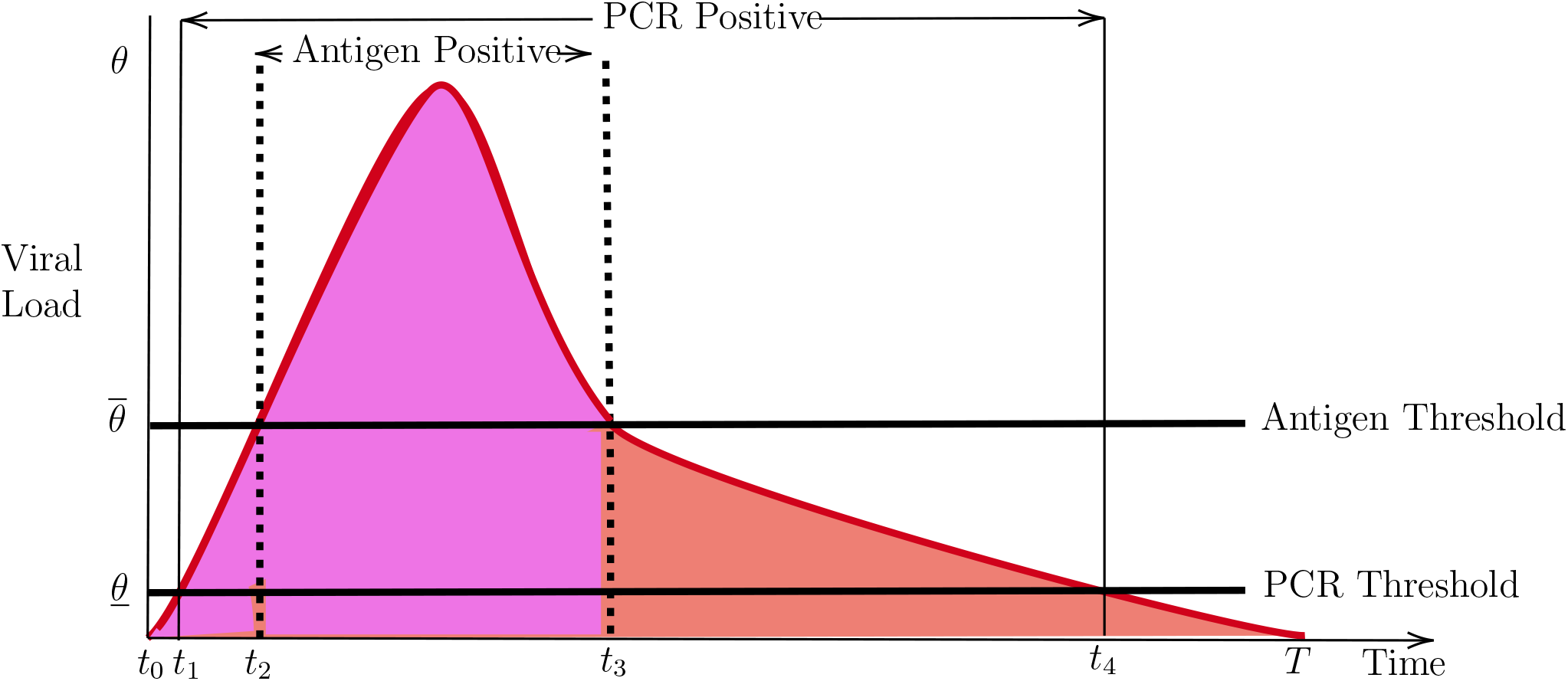
Sensitivity with Pre-Infectiousness

In this paper, I make a stronger claim: That even in the absence of a cost advantage or more frequent testing, a test with a higher limit of detection (e.g., an antigen test) may be more informative than a test with a lower limit of detection such as the ‘gold-standard’

PCR test. In particular, when a test’s efficacy is measured with respect to the decision being taken (isolation versus treatment), an antigen test can be more efficacious. In other words, it may not be ‘poorer’ but superior.

The outline for the paper is as follows. In Section 2, I provide a discussion of how tests are typically scored by regulators (using sensitivity and specificity) and also a review of the economic literature on testing. Section 3 introduces the model which involves a decisionmaker choosing actions of treatment or isolation based on potential costs of a utility loss from isolation, misdiagnosed treatment or broader contagion. Section 4 then examines how to construct sensitivity measures depending on the decision-type and how this relates to the information value of a test. Section 5 considers an extension to take into account pre-symptomatic screening for infection. A final section concludes.

## 2 Test Scoring

The primary means of scoring tests for clinical purposes is to calculate their *sensitivity* (that is, the probability that an individual with a condition tests positive for that condition) and *specificity* (that is, the probability that an individual without that condition tests negative for that condition). These have their analogues in Type I and Type II errors with sensitivity measuring the lack of false negatives and specificity the lack of false positives. Consequently, depending on test parameters, a test designer often faces a trade-off between test sensitivity and specificity.

These measures are used to score the efficacy of tests. A PCR test for SARS-CoV-2 typically has a specificity of 99% and a sensitivity between 80-98% depending on a number of factors including how skillfully a practitioner is able to capture a sample from an individual. If the pre-test (or prior) probability that a patient is infected is 5%, a test with 90% sensitivity and 99% specificity will have a false negative rate of 1% (i.e., 1% of those who test negative are not negative) and a false positive rate of 17% (i.e., 17% of those who test positive are not positive). By contrast, an antigen test – which looks for particular chemicals associated with SARS-CoV-2 – has a specificity equivalent to PCR tests but a potentially much lower sensitivity (as low as 84-97% compared with the best practice RT-PCR);^5^ implying that many, who are actually infected, will test negative for the coronavirus. However, it is important to note that (i) non-PCR tests have their sensitivity and specificity measured compared to PCR tests and (ii) PCR tests define their sensitivity and specificity with respect to infection, not infectiousness.

The terms sensitivity and specificity were coined by Yerushalmy (1947) who was examining the decision-theoretic foundations of using X-rays to inform on diagnosis. Sensitivity was the “probability of correct diagnosis of ‘positive’ cases” and specificity was the “probability of correct diagnosis of ‘negative’ cases.” In each case, the measure was tied to the purpose of the diagnosis. With virus detection, the purpose of a test is to inform a treatment decision in which case the diagnosis is whether an individual is infected or not. By contrast, with virus mitigation, the purpose of a test is to inform an isolation or quarantine decision in which case the diagnosis is whether an individual is infectious (or contagious) or not. Because the decisions are different, so too should be the measures of sensitivity and specificity even if the underlying target is similar at a molecular level.

In the US, all clinical tests are regulated by the Federal Food and Drug Administration (FDA). When approving a test for clinical purpose this is done with regard to its usefulness in treatment. Thus, PCR tests and antigen tests are scored on the same criteria. However, as will be demonstrated, this score is misleading when the purpose of a test is for a pandemic mitigation rather than a treatment decision. For such decisions, you want to diagnose an individual as infectious or not, rather than infected or not. A test that is less sensitive for infection may be more sensitive with regard to infectiousness.

It is important to note that PCR tests can provide information that can indicate infectiousness rather than infection. As mentioned before, the number of cycles a PCR test has to go through before rendering a positive result is a measure of the viral load in an individual. This cycle count (or Ct measure) is part of any PCR test. However, the reporting of the test results is usually a binary “positive” or ‘negative” outcome that discards this information. Some epidemiologists have called for a reporting of the Ct result as a matter of course (Tom and Mina (2020)).^6^ In the US, labs are not legally allowed to report Ct numbers so results are binary as a matter of regulation.^7^

Thusfar, the economics literature has focused on other issues regarding testing. Notably, Galeotti et al. (2020) do provide an exposition of taking an information theoretic approach to the value of testing but do not raise issues of infectiousness (as opposed to infection). Other work that examines the informational value of testing examines how to allocate costly or scarce tasks on the basis of available data or observations that underpins pre-test probabilities (see Ely et al. (2020) and Kasy and Teytelboym (2020)). Bergstrom et al. (2020) examine the optimal frequency of testing to reduce contagion. Finally, there is a literature on the impact widespread testing might have for behavioural choices of economic agents (Eichenbaum et al. (2020); Deb et al. (2020); Acemoglu et al. (2020); Taylor (2020) and Gans (2020)). This present paper is the first that examines the particular issues that arise from testing for infectiousness in an information-theoretic way.

## 3 Model Setup

The decision-maker (DM) is a public health authority who chooses two actions: a treatment action, *d_i_* = 0 (no treatment) and *d_i_* = 1 (treatment), and an isolation action, *a_i_* =1 (don’t isolate) and *a_i_* = 0 (isolate) for each individual 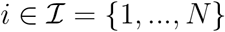 with a payoff of:

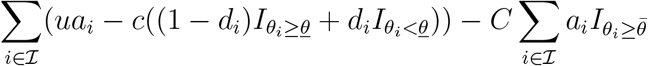

where *θ_i_* ∊ [0,1] is a measure of the viral load of an individual and I is an indicator function that takes a value of 1 if the condition is met and 0 otherwise.^8^ This state is not known to the DM. In contrast to Ely et al. (2020) the state, *θ_i_*, is not binary but is a continuum with distribution function, *F*(9) that is common for all individuals. There is a mass point at *F*(0) that could be interpreted as the share of the population who are not infected with the virus.

In this setup, *u >* 0 is the utility of a non-isolated agent *i*, *c* is the individual cost of a mistreatment^9^ and *C* is the social cost of not isolating an infected individual.^10^ Thus, 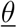 is the threshold for the viral load, above which an individual is considered to be *infected* with the virus and can benefit from treatment. By contrast 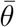 is the threshold for the viral load, above which an individual is considered to be *infectious*.

### 3.1 Perfect information

If the DM had perfect information regarding 9_i_, they would choose *d_i_* = 1 if and only if 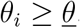. With respect to the isolation decision, for 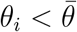, DM chooses *a_i_* = 1; and for 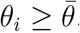, they choose *a_i_* = 0 if *u* ≤ *C* and *a_i_* = 1 otherwise. It will be assumed that *u* ≤ *C* always holds so that isolation is the optimal choice if the viral load is above the infectiousness threshold.

### 3.2 No test

By contrast, suppose that the DM had no information regarding any individual’s 9_i_. What choice would be optimal? Beginning with the treatment decision, note that it is assumed that the costs associated with a misdiagnosis are c regardless of the ‘direction’ of the error. Thus, for *i*, *d_i_* = 1 has an expected payoff of 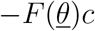 while *d_i_* = 0 has an expected payoff of 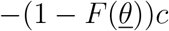. Thus, DM will treat rather than not treat if:

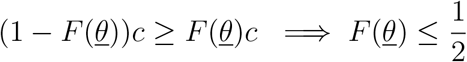

That is, blanket treatments are optimal if prevalence 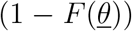 is high. For the isolation decision, the payoff from *a_i_* = 0 (isolation) for all *i* is (by our normalisation) 0 while the expected payoff if *a_i_* = 1 for all *i* is: 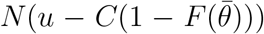. Thus, isolation is an optimal decision if:

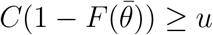

Here, high numbers of infectious individuals 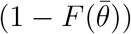 triggers a blanket isolation or lockdown decision.

## 4 Test Sensitivity

Suppose that there exists a test that can be deployed that will detect viral load above a certain point 9. In other words, the signal, s_i_ provided by a test is binary with ‘+’ if *θ_i_* ≥ *θ* and ‘−’ otherwise. Thus, if you conduct a test on an individual i, then with probability 1 − *F*(*θ*) it will return a positive result and with probability *F*(*θ*) a negative result.

### 4.1 Sensitivity of a test for infection

As noted in Section 2, regulators score the efficacy of clinical tests but measuring the sensitivity and specificity of those tests. However, these tests must be conducted with respect to the decision being taken and, for regulators, this is often for the purpose of informing treatment interventions (i.e., diagnosis). Thus, a test for the presence of a virus would provide information as to whether someone was infected and in need of potential treatment. This means that sensitivity and specificity would be considered with respect to 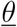. In medical terms this means that, prior to a test, the pre-test probability (or prior) that someone is infected is 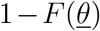; likely the population level of prevalence. In the case of the test described above, specificity 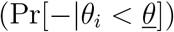) and sensitivity 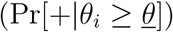) are:

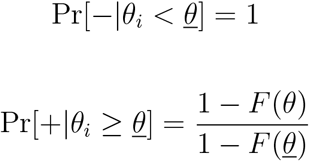

These are stated on the assumption that 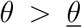. This is a reasonable assumption. For instance, for Covid-19, 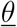 is often considered to be close 0. Under this assumption, if a patient is not infected, then they test negative for sure and so the specificity of the test is 100 percent. However, sensitivity is less than 100 percent because a negative test does not imply that the individual is negative. Note that specificity collapses to 1 as 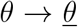 because 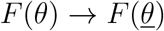.

What does the treatment decision look like with a test of *θ*? If the test is positive, the probability that you are positive is 1. If the test is negative, the probability that you are positive is:

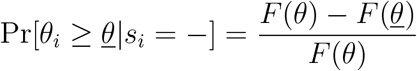

and the probability that you are negative is:

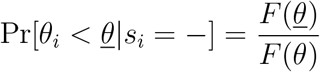

Thus, the DM will decide to not treat rather than treat on the basis of a negative test if:

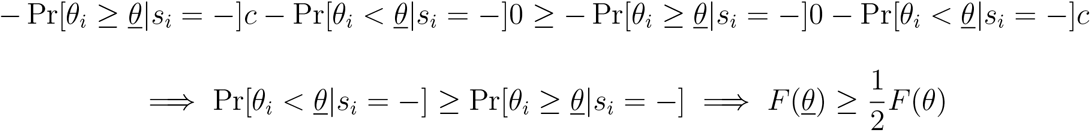

Note the critical role of 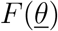, the pre-probability that someone is not infected, in this decision. The higher is 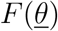 (i.e., the lower expected prevalence is), the more likely a negative test will trigger a decision not to treat the individual. In other words, with an imperfect diagnosis test, the DM will hold back on treatment somewhat for imperfect tests. This highlights the importance of obtaining more information regarding the likelihood of infection for an individual prior to interpreting test results (e.g., by observing for symptoms or having a recent other test).

What is the overall value of a test, *θ*, relative to not performing a test? Note, first, that if 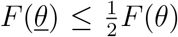, then treatment is a dominant action for the DM and will be chosen regardless of the signal. Thus, the test has no value. If 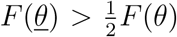, then the treatment action matches the test result. DM’s expected payoff prior to administering the test is:

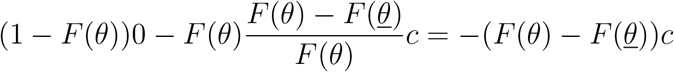

By contrast, if no test is administered, DM’s expected payoff is 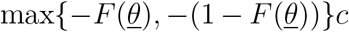. This means that the value of a test, v(9), is:

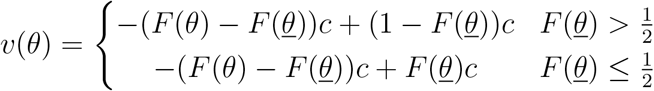

How do these relate to sensitivity? Let 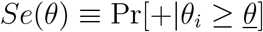. Then 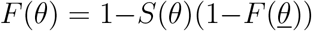. Substituting this into the value of a test we have:

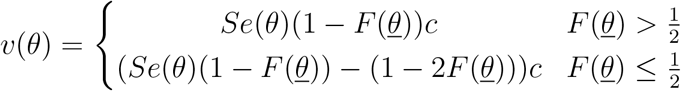

Thus, a test is of most value if sensitivity, *Se*(*θ*), and prevalence, 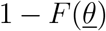 are both high up to a point where 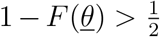. Beyond this point, the default action without a test switches to treatment and, thus, the value of a test is reduced.

### 4.2 Sensitivity of a test for infectiousness

One potential way of controlling the spread of a virus is to test in order to find infectious people and isolate them. While a test for infectiousness will likely look for the similar viral markers as a test for infection, there is evidence that infectiousness is critically dependent on the viral load (Tom and Mina (2020)). Thus, the threshold for whether someone is infectious is higher than that for whether they are infected. This is captured in the assumption that 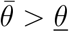. Here we examine how this impacts on the measurement of sensitivity and specificity.

The first thing to note is that the pre-test probability that someone is infectious is 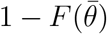 which is lower than the pre-test probability that someone is infected. For a test of infectiousness, the specificity 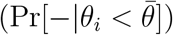 and sensitivity 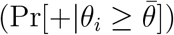 are:

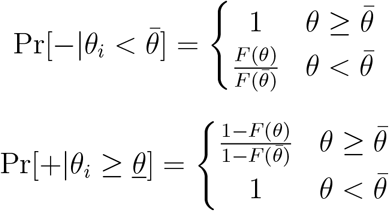

This demonstrates something very interesting. The monotonicity of the measures of sensitivity and specificity in *θ* are contingent on *θ* being above the threshold for an intervention.

While this was arguably a reasonable assumption for testing whether someone was infected with a virus, it is less obvious for whether someone is infectious or not. Indeed, as discussed in the introduction, many of the standard (and, indeed ‘gold-standard’) tests for Covid-19 were likely to detect the presence of the virus in very small concentrations. By contrast, infectiousness relies on the virus have a high concentration in an individual and, hence, those standard tests will detect the virus at levels well below 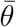; the threshold at which someone is said to be infectious. In this case, the test can return a positive result even where 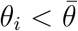 generating a false positive with respect to infectiousness. Thus, while a test with 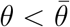 has 100 percent sensitivity, as *θ* falls, the specificity of the test falls implying that a DM would make more errors from false positives – i.e., isolating individuals who should not be isolated and incurring an utility loss of u each time.

Given this, how will the DM use the information from these tests to inform their isolation decision? Let’s consider a test with 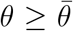 first. In this case, a positive test means you are infectious with probability 1. For a negative test,

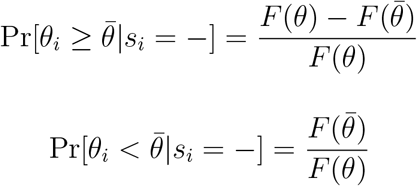

Thus, the DM would choose not to isolate an individual with a negative test if 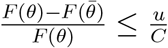. If this condition did not hold, the test would have no value at that time. Given this, if the test has value, DM’s expected payoff from administering the test is:

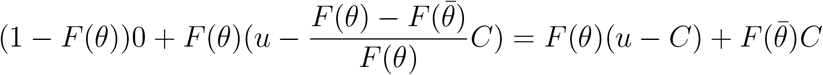

If no test is administered, DM’s payoff is 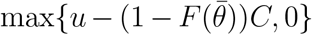. Thus, the value of a test for infectiousness, *V*(*θ*) is:

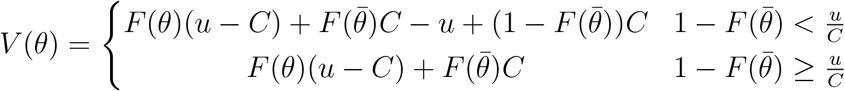

We can consider how these relate to sensitivity by letting 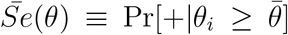. Then 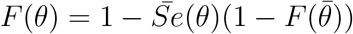. Substituting this into the value of a test we have:

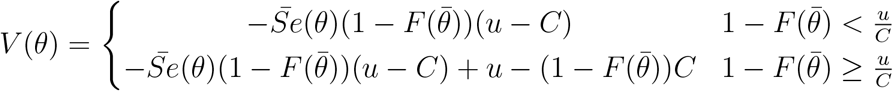

This is increasing in 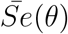 by our earlier assumption that *u* ≤ *C*.

Now, consider the case where 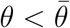. In this case, a negative test means *i* is not infectious with probability 1 as sensitivity is equal to 100 percent. For a positive test,

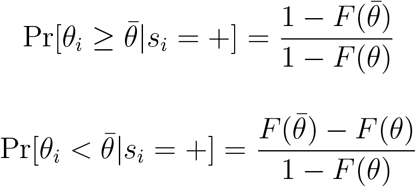

Thus, the DM would choose to isolate an individual with a positive test if 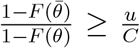. If this condition did not hold, the test would have no value. Given this, if the test has value, DM’s expected payoff from administering the test is:

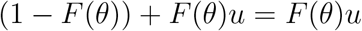

If no test is administered, DM’s payoff is 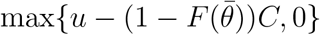.

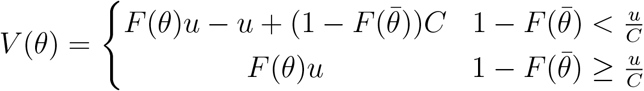

We can consider how these relate to specificity by letting 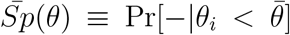. Then 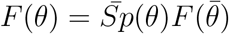. Substituting this into the value of a test we have:

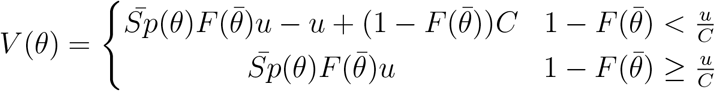

This is increasing in 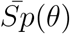.

### 4.3 The optimal test for infectiousness

It has been demonstrated above that the value of a test for infection, υ(*θ*) is decreasing in *θ* until 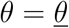. By contrast, let’s examine the impact of 9 on a test for infectiousness.

#### Proposition 1

*V*(*θ*) *is increasing in θ for* 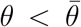 *and decreasing in θ for* 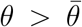 *with a maximum at* 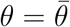.

##### Proof

When 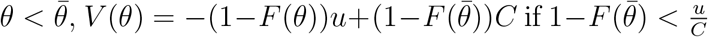 and 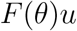 otherwise. In each case, 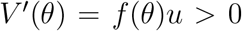. When 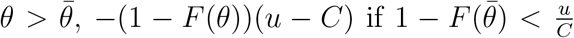 and 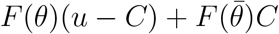 otherwise. In each case, *V*′(*θ*) = *f*(*θ*)(*u* − *C*) < 0. ■

Figure 2 plots *V*(*θ*) is a function of 9 for the cases where, the current share of infectious agents, 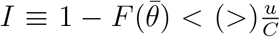. Because *F*(0) > 0, each starts at a positive value at *θ* = 0, rises until 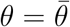 and falls thereafter.

This is the main result of the paper. When tests are scored on the basis of sensitivity with regard to infection (for the purposes of a treatment decision), these favour tests with a lower *θ*. However, when these tests are below 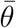, the threshold for infectiousness, requiring a lower *θ reduces* the value of those tests. This result arises even though we have not taken into account the cost of tests, where a test cost is likely to be higher the lower is *θ*, nor their frequency. In other words, scoring tests for infectiousness on the basis of sensitivity of tests for infection, leads to *less* informative tests for infectiousness and hence, would end up isolating too many individuals. This would be economically wasteful.

## 5 Pre-infectiousness

The above analysis assumes that when 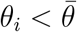, the optimal decision is to not isolate *i*. For a virus like SARS-CoV2, the viral load only rises above 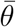, if at all, after three or so days from the point the individual becomes infected. Unless tests are being conducted very frequently – of the order of every 1-2 days around the time a person becomes infected – it would also be optimal to isolate someone with a low viral load who has just been infected. Thus, examining whether 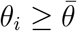 or not is insufficient to obtain the optimal decision.

While frequent testing can overcome this difficulty, here I want to note how to adjust the sensitivity of a test for infectiousness to take this into account. Figure 3 shows a typical path for the viral load and compares a (perfect) PCR test for infection (i.e., 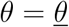) with a (perfect) Antigen test for infectiousness (i.e., 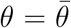). In this figure, the optimal decision is to isolate the patient from period t_0_ to t_3_. If 1 − *F*(0) is the probability that an individual carries some amount of the virus, then the probability that they test negative for an antigen test with a threshold of 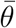 is 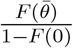 which is a false result with probability 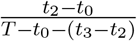. By contrast, a negative PCR test, which happens with probability 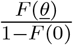 is false for infectiousness with probability 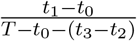.

Given this, the specificity and sensitivity of the PCR test for infectiousness is:

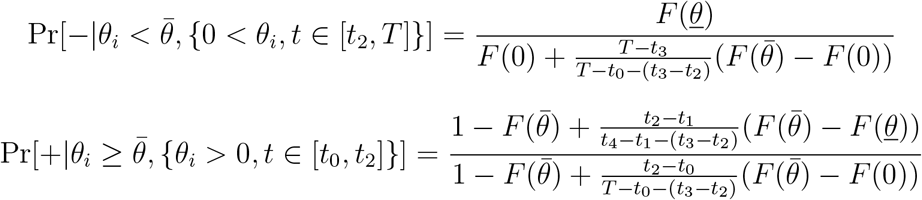

The difference between these measures and those provided earlier arises due to the recognition of potential infectiousness between *t*_0_ and *t*_2_. When this gap disappears, these measures converge to the earlier ones for the case where 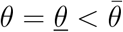.

For the antigen test, the specificity and sensitivity for infectiousness become:

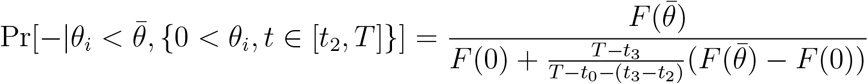

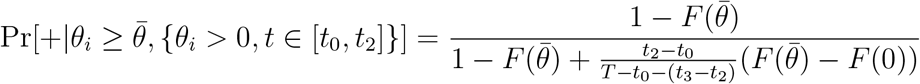

Comparing this with the measures for the PCR test, the antigen test still has higher specificity but the ranking on sensitivity becomes less clear cut. The PCR test risks false positives, as they did before, of people who have already been infectious but are still infected but picks up, in a way that the antigen test does not, the pre-infectious but infected individuals (that is, 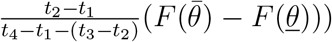. In particular, the antigen test, even with 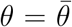 is less than 100 percent sensitive because of the presence of pre-infectious individuals.

This adjustment does not alter the broad conclusion of Proposition 1 that a test for infectiousness should not require a threshold *θ* to be as low as possible. It does suggest that the optimal test may involve 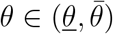. These analyses presume that an infected individual receives at most one test while they are infected. Of course, if the tests were conducted more frequently (something possible with cheaper antigen tests that have immediate results), then the information they provided together could be used to form a clearer picture of where in the viral life-cycle an infected individual was.

## 6 Conclusion

This paper has examined the consequences of choosing a test scoring method that does not match the decision being taken. If sensitivity standards for test of SARS-CoV-2 infection are used to score tests for infectiousness, the value of tests in informing an isolation decision is reduced. Insisting on treatment sensitivity requirements leads to more false positives in the isolation decision; that is, individuals are isolated even though they are not infectious. This similarly leads to other costs not modelled here. The decision to release someone from isolation is usually predicated on a negative test which, if made on the basis of infection, would cause people to be isolated for too long. Indeed, they are even safer given that they have previously been infectious. In contact tracing, a positive PCR test is used to inform a costly exercise in tracking down contacts and isolating them. It is likely that most of those efforts are wasted unless those decisions are informed by a test more suited for infectiousness or, alternatively, using the viral load (or Ct) information in the PCR test. Currently, that information is not collected or reported.

## Data Availability

No data is used in the paper.

## Footnotes

1 Sometimes people look to rank tests according to the Blackwell (1953) criteria of informativeness. Here, the tests I will examine do not naturally correspond to that ranking and so the focus is on the value of a test per se.

2 For example, Larremore et al. (2020) and Paltiel et al. (2020). Also, these tests require PPE for humans to administer, adding to the cost.

3 Source: Larremore et al. (2020)

4 Larremore et al. (2020) also point out that a test taken at Day 15 might be positive under the PCR test (e.g., point D) but, by that time, the virus itself is dead.

5 https://www.cdc.gov/coronavirus/2019-ncov/lab/resources/antigen-tests-guidelines.html Döhla et al. (2020) found antigen sensitivity compared with PCR of 36%.

6 This can be particularly useful if patients have multiple tests because the change in the Ct number can indicate where they are on the lifecycle of the virus.

7 My source for this is Michael Mini (a Harvard epidemiologist) who stated as such here https://youtube/3seIAs-73G8?t=3544 I have not been able to find the specific regulation, however.

8 A simplifying assumption here is that individuals are identical from the perspective of DM. This is innocuous unless there are situations where the DM has specific information about *i*’s utility that differs from others.

9 In reality, the cost of mistakenly treating someone and the cost of mistakenly not treating them are likely to be different. However, since the treatment decision is not the main focus of this paper, the losses are assumed to be symmetric for simplicity.

10 This is a simplification as the marginal cost of an additional infected person who is able to interact with others depends upon the number of susceptible people remaining in the population. However, using an more complex and epidemiologically founded cost model is unlikely to change the broad conclusions of this paper.

## Notes

### Competing Interest Statement

The authors have declared no competing interest.

### Funding Statement

No funding has been provided.

### Author Declarations

No oversight body required as paper is theoretical and does not involve human subjects.

